# Germline Allelic Expression of Genes at 17q22 Locus Associates with Risk of Breast Cancer

**DOI:** 10.1101/2021.12.10.21267625

**Authors:** Filipa Esteves, Joana M. Xavier, Anthony M. Ford, Cátia Rocha, Paul D.P. Pharoah, Carlos Caldas, Suet-Feung Chin, Ana-Teresa Maia

## Abstract

Translation of GWAS findings into preventive approaches is challenged by identifying the causal risk variants and understanding their biological mechanisms. We present a novel approach using AE ratios to perform quantitative case-control analysis to identify risk associations, causal regulatory variants, and target genes. Using the breast cancer risk locus 17q22 to validate this approach, we found a significant shift in the AE patterns of *STXBP4* (rs2628315) and *COX11* (rs17817901) in the normal breast tissue of cases and healthy controls. Preferential expression of the G-rs2628315 and A-rs17817901 alleles, more often observed in cases, was associated with an increased risk for breast cancer. Analysis of blood samples from cases and controls found a similar association. Furthermore, we identified two putative *cis*-regulatory variants – rs17817901 and rs8066588 – that affect a miRNA and a transcription factor binding site, respectively. Our work reveals the power of integrating AE data in cancer risk studies and presents a novel approach to identifying risk - case-control association analysis using AE ratios.

## Introduction

Genome-wide association studies (GWAS) identified hundreds of loci associated with breast cancer risk, which require functional characterization to reveal novel insights into the risk-associated pathophysiological mechanisms and lead to preventive treatments. The location of most GWAS signals within non-coding regions suggests a regulatory role for the risk-associated variants, which was confirmed for some loci ^1–3^. However, the high populational frequency of the risk-associated alleles and the complex linkage disequilibrium structure in the risk-loci precludes their identification and the target genes they regulate.

Identifying the target genes regulated by risk-variants usually includes physical interaction studies (e.g., chromatin conformation capture ^4^), *in-vitro* assays evaluating protein binding modification at regulatory elements (e.g., band shifts and transfection assays ^1^), and more integrative approaches ^5^, but often lack direct *in-vivo* validation in the complex genomic context of the disease’s tissue of origin. However, as these variants regulate gene expression in an allele-specific manner, their target genes can be detected by measuring and comparing allelic expression (AE) levels in heterozygous individuals for a transcribed variant ^6,7^. Compared to expression quantitative trait loci (eQTLs) analysis ^8–10^, analyzing AE ratios has the advantage of isolating cis-acting effects while controlling for *trans*-acting and environmental ones ^11–13^, of detecting epigenetic effects ^12,14^, and showing increased statistical power ^15^. Still, the informative value restriction to heterozygous samples and the complexity of data processing and analysis required have limited AE analysis.

Given this evidence, we propose integrating AE studies carried in the normal breast tissue in the functional characterization of known risk loci. Additionally, to test whether inherited allele-specific expression of target genes act as a mechanism of predisposition to breast cancer, we also propose to compare allele-specific expression levels between cases and controls. Allelic expression analysis was used before to identify predisposition to disease in a qualitative manner ^16,17^, but we propose to take advantage of its quantitative nature to increase the statistical power of a case-control comparison.

To evaluate the value of integrating AE analysis in the characterization of risk to breast cancer, we tested our approach in the study of the locus 17q22, which was associated with risk of breast cancer in three studies ^18–20^, including male breast cancer ^21^; and possibly with breast cancer survival ^22^. The first variant associated with risk was rs6504950 (intronic in *STXBP4*) (per-allele odd ratio OR = 0.95; 95%CI = [0.92; 0.97]; *P* = 1.4 × 10^−8^) ^18^, and more recently a fine mapping of the locus established rs2787486 (intronic in *STXBP4*) as the lead risk-SNP (OR = 0.92; 95%CI = [0.90; 0.94]; *P* = 8.96 × 10^−15^) ^20^. Whilst data indicates that these alleles modify disease risk target the genes *COX11* and *STXBP4* ^4,5,20^, they have not been functionally validated, and causal variants remain unidentified ^5^. In this work, we first analyzed AE patterns in the normal breast tissue of controls to identify the genes under the effect of *cis*-regulatory variants in the locus; then compared the distribution of AE ratios measured in the normal breast tissue of patients with that of controls to assess association with risk and identify target genes; next, we assessed if blood could be used as a surrogate tissue for BC risk assessment based on AE studies; and finally we mapped the regulatory variants driving the AE of *STXBP4* and *COX11*.

## Material & Methods

### Samples

All samples were collected for this study following written informed consent from all donors. All procedures followed were per the established rules of the Addenbrooke’s Hospital Local Research Ethics Committee (REC references 06/Q0108/221, 07/H0308/161, and 04/Q0108/21 for normal breast tissue from healthy controls, normal-matched tissue, and blood from breast cancer patients, respectively) and the Eastern Multicentre Research Ethics Committee (SEARCH Study) ^23^. The original studies provide the full description of the samples 23–25.

### Cell Lines

Lymphoblastoid cell lines derived from unrelated CEPH individuals (Coriell Cell Repository) were used as described previously ^24^. Breast cancer cell lines MCF7 and HCC1954 were cultured in DMEM and RPMI-1640 media, respectively, supplemented with 10% fetal bovine serum, 1,000 U/mL penicillin, 1 mg/mL streptomycin, and 2 mM L-glutamine (Gibco, Thermo Fisher Scientific). MCF7 and HCC1954 were cultured in DMEM and RPMI-1640 media, respectively, supplemented with 10% fetal bovine serum, 1,000 U/mL penicillin, 1 mg/mL streptomycin, and 2 mM L-glutamine (Gibco, Thermo Fisher Scientific).

### Nucleic Acid Preparation and Processing

Extraction of DNA and total RNA from all samples was previously described ^23–25^. cDNA was synthesized using the SuperScript™ First-Strand Synthesis System (Invitrogen), from 50 ng of total RNA and a mixture of oligo(dT)_20_ and random hexamers, according to the manufacturer’s instructions. Target-specific preamplification of cDNA was performed with TaqMan™ PreAmp Master Mix (2X) (Applied Biosystems), pooled TaqMan™ SNP Genotyping Assays (0.2X) (Applied Biosystems), and 1.25 μl of cDNA. Thermal cycling conditions consisted of enzyme activation at 95°C for 10 min, followed by 8 or 14 cycles of denaturation at 95°C for 15 sec and annealing/extension at 60°C for 4 min. Finally, products were diluted 1:5 before use in subsequent reactions.

### Genotyping

Genotyping was performed using TaqMan™ SNP Genotyping Assays (a custom assay for rs17817901 and predesigned assays C_15903698_10 and C_30379485_10, for rs2628315 and rs9899602, respectively), under cycling conditions per the manufacturer’s instructions. Reactions were prepared in a final volume of 5 μl with TaqMan™ Universal Master Mix II, with UNG (2X) (Applied Biosystems), TaqMan™ SNP Genotyping Assay (40X) (Applied Biosystems), DNase/RNase-free water, and 8 ng of DNA. Reactions were performed in a Bio-Rad CFX384 system (Bio-Rad).

### Allelic Expression (AE) Analysis

Using previously generated data from a genome-wide microarray study ^26^, we quantified AE as described previously ^27^. Briefly, we identified the transcribed SNPs where AE could be measured (aeSNPs) in the risk locus included in the data and extracted the AE levels for all heterozygous individuals at each aeSNP. We calculated the normalized AE ratios (AE ratios_norm) as the log2 [(expression of alternative allele) / (expression of reference allele)] normalized by the same ratio calculated from genomic DNA data (gDNA) to account for copy number variation and correct for technical biases. To test if the distribution’s mean of the AE ratios_norm for each aeSNP was equal to zero (null hypothesis), we used a one-sample two-sided Student’s t-test. P-value was corrected for multiple testing using the Benjamini & Hochberg correction from q.value R package and considered significant when FDR was lower than 1%.

Allele-specific expression was also quantified using real-time PCR for the case-control association study, with the TaqMan™ SNP Genotyping Assays indicated above and cDNA, and as described previously ^24^. Experiments were performed on 96.96 Dynamic Arrays™ IFC in the Biomark™ HD system (Fluidigm) and on a CFX384 real-time PCR machine (BioRad). Here, AE ratios were calculated as the log2 [(alternative allele) / (reference allele)], without normalization. Standard curves consisting of serial dilutions of DNA from CEPH lymphoblastoid cell lines heterozygous for each SNP were used to determine the quantitative performance. Cases, controls, standard curves, and at least two no-template controls (NTC) were analyzed in triplicates simultaneously in each experiment, and cases and controls were solely compared within each experiment.

### Case-Control Association Analysis

To detect AE ratios associated with a risk of breast cancer, we calculated the effect size, given with magnitude and direction of the difference, between the AE ratios of cases and controls, in breast tissue and blood. For this purpose, we used the Hedges’ *g* (effect size) test, which is a standardized mean difference method that normalizes for sample size, particularly suitable for small ones (< 20 samples in each group). More specifically, each test included taking 5000 bootstrap samples; the confidence interval is bias-corrected and accelerated. We also report the p-value(s) for the likelihood(s) of observing the effect size(s) if the null hypothesis of zero difference is true, assessed by the Mann-Whitney test. P-values were permuted with 5000 label reshuffles of the controls and cases. All Cumming and Gardner-Altman estimation plots, and statistical tests were performed using the online tool available at www.estimationstats.com ^28^. Data for the multiple experiments run for this analysis can be found in the GitHub repository indicated in “Data and Code Availability”.

### Mapping of Candidate Regulatory Variants

Annotation of aeSNPs was performed using the R package Biomart ^29^ and Ensembl Browser [https://www.ensembl.org/index.html]. LD between rs760482 and each aeSNP was retrieved using ensemblr R package [https://github.com/ramiromagno/ensemblr] and the 1000 Genomes project Phase3 European population data. aeSNP with mean AE ratios significantly different from zero were identified using a two-sided t-test for unequal variances. P-value was corrected for multiple testing using the Benjamini & Hochberg correction in the q.value R package and considered significant when FDR was lower than 1% ^30^.

### *In-Silico* Functional Analysis of Variants

Candidate regulatory SNPs (rSNPs) were retrieved using SNP Annotation and Proxy Search (SNAP v2.2 ^31^ and the HaploReg v4.1 ^32^ tools, based on high linkage disequilibrium (LD, r^2^ ≥ 0.8 in the European population - CEU) with each daeSNP identified in the *locus*. Each candidate rSNP was assessed for a potential change of transcription factor (TF) binding at genomic regulatory regions, overlap with *DNase*I hypersensitivity sites, histone modifications (H3K4me1, H3K4me3, H3K27ac, and H3K9ac), and protein binding (ChIP-Seq data) on data from Roadmap Epigenomics and ENCODE projects ^33,34^. Epigenetic data and allele-specific position weight matrix (PWM) were also retrieved for human mammary epithelial cells (HMECs), breast myoepithelial primary cells, human mammary fibroblasts (HMFs), and MCF7 and T47D breast cancer cell lines, using HaploReg, RegulomeDB v1.1 ^32,35^. Candidate rSNPs with the potential to disrupt/affect miRNA binding were selected based on predicted allele-specific miRNA targeting, as previously described ^27^.

### Electrophoretic Mobility Shift Assay (EMSA)

EMSAs were carried out using the LightShift Chemiluminescent EMSA Kit (Thermo Fisher Scientific). Briefly, all binding reactions were performed in a final volume of 20 μL, including 20 mM Hepes, pH 7.4, 5% glycerol, 10 ng/μL Poly (dI-dC), 1X protease inhibitor, 1 mM DTT, 5 μ g of nuclear protein extract, and 60 fmol of the labeled oligonucleotide probe. We prepared nuclear protein extracts from MCF7 and HCC1954 cells using the NE-PER nuclear and cytoplasmic extraction kit (Thermo Fisher Scientific). Oligonucleotide sequences used in the assays included the reference and the alternative alleles and are listed in Table S1. Competitor unlabeled oligonucleotides were used at 10-, 33-, and 100-fold molar excess. Each EMSA was repeated at least twice for all combinations of nuclear protein extract and oligonucleotide.

### Luciferase Reporter Assay

We performed reporter assays using long oligonucleotides (80 bases) synthesized with three tandem copies of the hsa-miR-194-5p binding site (with either allele of rs17817901, Table S1) and internal restriction site of *BamHI*. These oligonucleotides were cloned into the pmirGLO Dual-Luciferase miRNA Target Expression Vector (Promega). MCF7 cells were transfected or co-transfected with an empty vector (control), vector with the cloned constructs, and synthetic miRNAs mimics (miRIDIAN microRNA Human hsa-miR-194-5p - Mimic, C-300642-03-0002, and miRIDIAN microRNA Mimic Negative Control #1, CN-001000-01-05, Dharmacon) using the DharmaFECT DUO Reagent (Dharmacon). Three replicates were included in each transfection. Twenty-four hours after transfection, luciferase assays were performed with the Dual-GLO Luciferase Assay System (Promega). Normalized firefly luciferase activity (firefly luciferase activity / *Renilla* luciferase activity) was determined and compared with the control for each construct. Pairwise differences in the mean of the tested conditions were assessed using Welch’s t-test.

## Results

### Genetic variants regulate genes in 17q22 risk-locus in strong linkage disequilibrium with the lead risk-SNP

Firstly, we assessed whether genes flanking (500Kb up and downstream) the GWAS lead SNP rs2787486 were under the control of *cis*-regulatory variants in normal breast tissue. For this, we calculated normalized allelic expression (AE) ratios at 29 aeSNPs located in *COX11, TOM1L1, STXBP4, HLF* and *MMD* (Figure S1). Twenty (69%) aeSNPs showed significant deviations from equimolar AE and were designated differentially allelic expressed SNPs – daeSNPs (Table S2). The observed differences between alleles reached a maximum of 13-fold at rs9303360. We identified daeSNPs in all genes in this region, supporting that all are targets of *cis*-regulatory variation.

The patterns of the AE ratio distributions are indicative of the linkage disequilibrium (LD) between the regulatory variant and the transcribed variant where AE is measured ^36^. Hence, to test a link between *cis*-regulation and risk, we examined the pairwise LD between the daeSNPs and the locus lead risk-SNP rs2787486 and matched it to the AE ratio distribution patterns. Four daeSNPs (located at *TOM1L1, COX11* and *STXPB4* genes) showed marked preferential expression of the same allele in all heterozygous individuals tested (Figure S1) and were in high LD (r^2^ ≥ 0.5) with rs2787486 (Table S2), suggesting that the same variant could confer risk and regulate gene expression levels.

One of these four daeSNPs, rs2628315, is in almost complete LD (r^2^ = 0.99) with the risk-variant and maps exclusively to the *STXBP4* gene (Table S2). At this variant, the allele preferentially expressed is associated with protection against breast cancer (Figure 1), suggesting that a higher expression of the A allele is beneficial. Concordantly, the GTEx project reports rs2628315 as an eQTL (expression quantitative trait locus) for *STXBP4* expression in mammary tissue (*P* = 9.68 × 10^−7^) ^37^.

**Figure 1.**
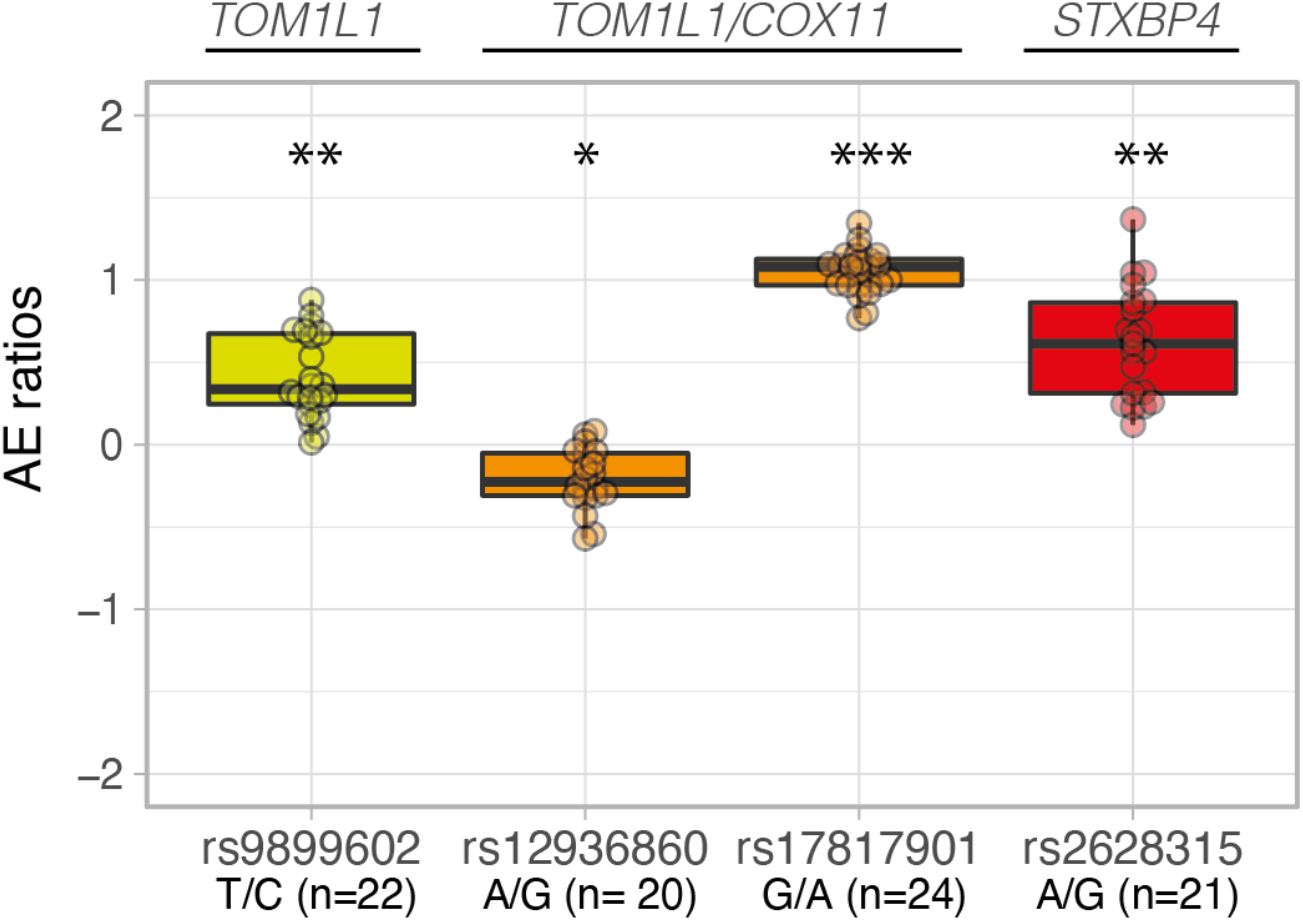
Genes in the 17q22 locus are under the effect of cis-regulatory variants genetically related to breast cancer risk variants. Boxplots of the allelic expression (AE) ratios for four variants in strong LD with lead risk-SNP rs2787486, located in the genes indicated above the graph. One-sample t-test for mean allelic expression equal to zero: * P < 10^−2^; ** P < 10^−5^; *** P < 10^−10^; Boxplots and data points are colored based on LD with rs2787486: yellow r^2^ >0.4, orange r^2^ > 0.50, and red r^2^ > 0.8.

Another two daeSNPs, rs12936860 and rs17817901, were in strong LD with rs2787486 (r2 = 0.57 for both, Table S2). These map to a shared region between *TOM1L1* and *COX11* genes but, rs17817901 showed the most significant differential AE pattern, as all heterozygotes preferentially expressed the alternative G allele. This AE ratio distribution is consistent with the daeSNP being in complete LD with the *cis*-regulatory variant (rSNP), creating the allelic effect ^36^, which facilitates the mapping of the latter (Figure 1).

Additionally, our results suggest that the preferential expression of the alternative G allele could correlate with the protective effect of the alternative C allele of rs2787486.

The fourth daeSNP in high LD with risk-variant rs2787486 is rs9899602 (r^2^ = 0.53) that maps exclusively to *TOM1L1*. It showed preferential expression of the reference T allele, which is correlated to the risk-associated A allele of rs2787486 (Figure 1).

These results suggest that the differential AE detected in all three genes could be associated with the risk of breast cancer and that all genes are candidate targets for the risk detected in the locus.

### AE ratios in normal breast tissue and blood are associated with breast cancer risk

Next, we sought to discern between chance colocalization and a true association between AE ratios and risk. We hypothesized that if risk-causing variants are *cis-*regulating genes in the 17q22 locus, then the AE ratios they generate should have distinct distributions in patients (cases) and healthy individuals (controls). Hence, we used AE ratios measured in the normal breast as a quantitative phenotype to perform case-control association analysis.

We carried out this analysis for the three daeSNPs displaying the highest LD with the risk-associated variant, each localized in one of the genes in the locus. As rs12936860 and rs17817901 are in complete LD, we only analyzed rs17817901. Data for the multiple experiments performed for this analysis are available in Supplementary Data.

The daeSNP rs2628315, located in an intron of *STXBP4*, showed the largest effect size (*g* = −1.237; 95%CI = [−1.928; −0.366]) (Figure 2), and the most significantly different AE ratio distributions (p.perm = 2 × 10^−4^). This result shows that the AE ratio distribution in the normal breast of cases is shifted towards the preferential expression of the reference G allele, the least expressed in controls. As rs2628315 and the risk-variant rs2787486 are in perfect LD, this result suggests that increased risk is associated with the preferential expression of the reference allele of both variants.

**Figure 2.**
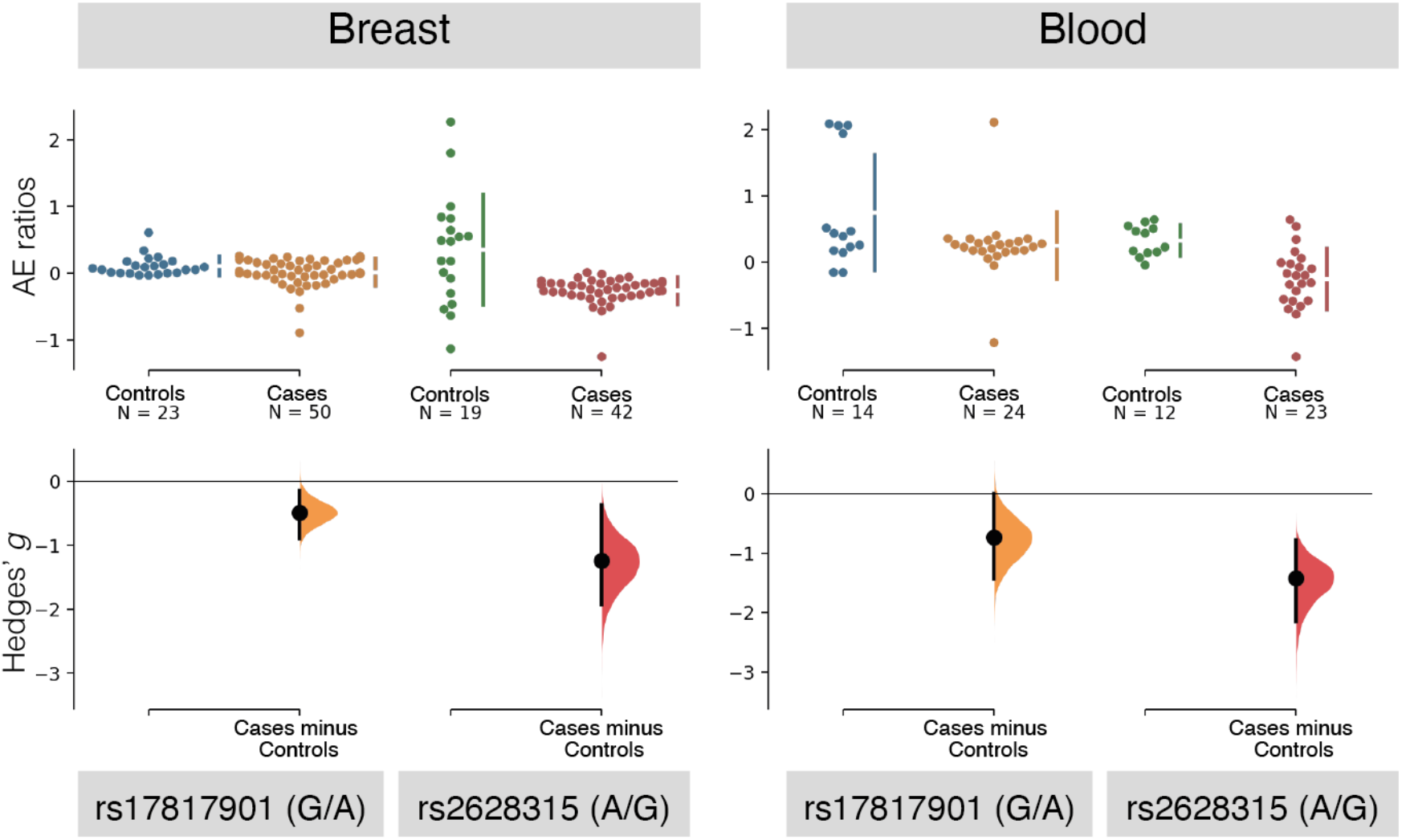
Case-control study using allelic expression ratios identifies risk in the 17q22 locus in breast tissue and blood samples. *Cumming estimation plot of Hedges’ g between breast cancer cases and controls for AE ratios calculated at rs17817901 (ratio calculated as allele G by allele A) and rs2628315 (ratio calculated as allele A by allele G) in normal breast and blood. The heterozygous individuals for each indicated variant and tissue are plotted on the upper axes, with controls displayed in blue and cases in orange* for rs17817901 and in green and red for rs2628315. *The vertical lines next to the raw data correspond to the conventional mean ± standard deviation error bars, where the mean of each group is indicated as a gap in the line. The Hedges’ g is plotted on the lower axes as a bootstrap sampling distribution (bootstrap n = 5000). The Hedges’ g values are depicted as dots, and the 95% confidence intervals are indicated by the ends of the vertical error bars*.

The analysis of rs17817901 also revealed a shift in the distribution of AE ratios in cases towards the preferential expression of the reference A allele with an estimated effect size of *g* = −0.486 (95%CI = [−0.889; −0.147]; p.perm = 5.34 × 10^−2^) (Figure 2). As rs17817901 is in strong LD with the risk-variant rs2787486, our results suggest that risk could be associated with a higher expression of the reference A allele of rs17817901. However, as rs17817901 locates in a genomic region shared by the *TOM1L1* and *COX11* genes, we considered both genes as candidate target genes for breast cancer.

However, the analysis of the daeSNP rs9899602 did not reveal any significant difference between the two populations (*g* = 0.038; 95%CI = [−0.593; 0.756]; p.perm = 0.89), suggesting that *TOM1L1* might not be a target gene for the risk detected via the lead-SNP rs2787486 (Figure S2). We note that rs9899602 is the daeSNP in weaker LD with the lead risk-SNP amongst those with significant DAE (Table S2). Integrated with the results obtained for rs17817901, this suggests that *COX11* is the most likely candidate among the two overlapping genes.

Based on the shared *cis*-regulation of breast cancer genes between breast and blood tissue ^12^, we next examined the associations described above in blood samples from cases and controls. For the daeSNP rs2628315, we found a comparable effect size (*g* = −1.419; 95%CI = [−2.156; −0.780]; p.perm = 0) and a significant difference in the AE ratio distributions of the two groups, with a concordant shift direction with that observed in breast tissue: preferential expression of the risk-associated G allele of rs2628315.

For the daeSNPs rs17817901, we found a larger effect size than was observed in breast tissue (*g* = −0.737; 95%CI = [−1.431; −0.002]; p.perm = 3.40 × 10^−2^), and in concordant direction – cases preferentially expressed the A-rs17817901 allele which is in strong LD with the risk-associated A-rs2787486 allele.

### Functional analysis reveals two rSNPs in the locus

Having found an association between the AE measured at rs17817901 and rs2628315 and the risk for breast cancer, we next aimed at pinpointing the regulatory variant(s) (rSNPs) responsible for this association. The distribution of the normalized AE ratios measured in breast tissue at rs17817901 (Figure 1) strongly suggests that the rSNP(s) generating this effect is(are) in strong to perfect LD with rs17817901 ^36^. Therefore, 106 variants in strong LD (r^2^ ≥ 0.8) with rs17817901 were analyzed *in-silico* for known functional data and predictions of functionality, with the strongest candidates subsequently tested *in-vitro* (Table S3). These analyses identified four candidate rSNPs - rs17817901, rs8066588, rs9896044 and rs9891865-regulating the binding of transcription factors and a miRNA.

Data for rs17817901 from public databases indicated limited evidence for functionality on breast tissue. However, it showed that the variant overlaps an enhancer element active in T cells and is an eQTL for all three genes in the locus in various tissues (not breast or blood) (Table S3). Furthermore, rs17817901 maps to the shared 3’UTRs of the genes *TOM1L1* and *COX11*, and we predicted before that its alternative G allele generates a binding site for hsa-miR-194-5p (context score = −0.229) ^27^, an oncogenic miRNA expressed in breast ^38–40^. This prediction was validated in reporter assays using a mimic oligo of the oncogenic hsa-miR-194-5p, which showed decreased reporter activity for the alternative G allele (protective) compared to the A allele and the empty vector (Figure 3A). A comparable difference was observed for endogenous levels of the miRNA, although non-significant.

**Figure 3.**
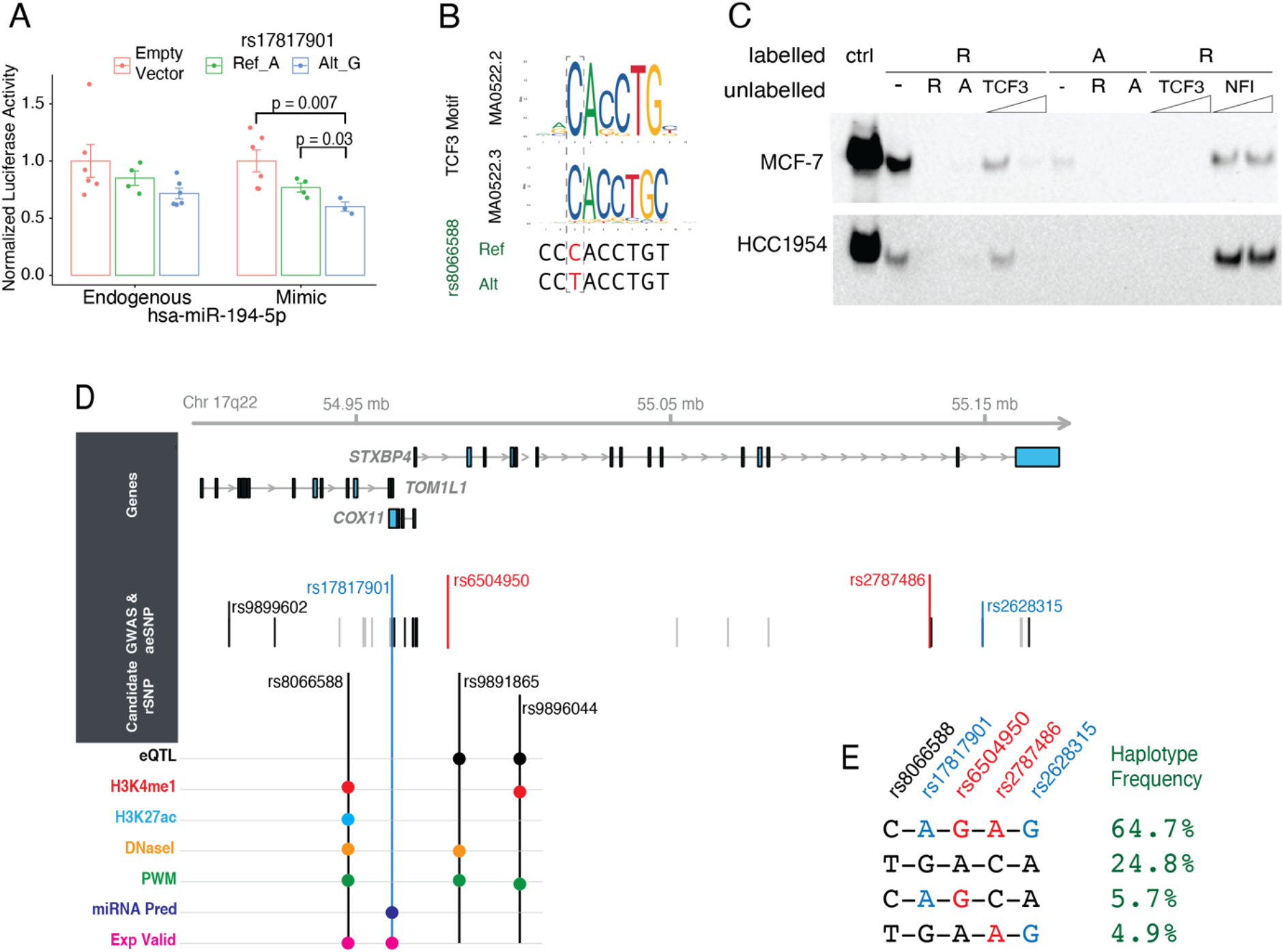
Functional characterization of candidate rSNPs related to breast cancer risk variants in the 17q22 locus. **A** - Luciferase assays showed a more significant reduction of signal for the alternative G allele of rs17817901, both under endogenous miRNA conditions and using a mimic oligo of hsa-miR-194-5p (y-axis shows the normalized luciferase Firefly/Renilla ratio) p-values indicated correspond to Welch’s test). **B** - The reference C allele of rs8066588 is predicted to generate a motif for the binding of TCF3, as indicated by the two consensus motifs indicated (MAO522.3 and MAO522.2). **C** - EMSA experiments using protein extracts from MCF-7 and HCC1954 cell lines show preferential binding of the reference C allele of rs8066588 (R – reference, A – alternative alleles); competition with oligo of known binding site forTCF3 competes with observed binding, which does not occur with negative control oligo (NFI binding motif). **D-** Genomic landscape of 17q22 locus showing the RefSeq genes in the top panel; then the location of the variants in which AE ratios were measured (aeSNPs), with black indicating daeSNPs with significant differential AE, in blue the risk-daeSNPs and in red the GWAS-SNPs; next is the location of candidate regulatory variants rSNPs and below the epigenetic, molecular and experimental evidence (eQTL, histone modifications, DNaseI hypersensitivity sites, PWM position weight matrix, miRNA prediction, and experimental validation). **E** – Haplotypes constructed with genotyping data of normal breast samples from controls included in this study, with the frequency indicated. The color scheme is as above, with risk-associated alleles for daeSNPs and GWAS-SNPs.

Next, the candidate rSNP rs8066588, an intronic variant to *TOM1L1*, is in complete LD with rs17817901 and strong LD with the risk lead-SNP (r^2^ = 0.85). According to the GTEx project, this variant is also an eQTL for *STXBP4* and an sQTL (splicing quantitative trait locus) for *COX11* in breast tissue. Also, we confirmed *in-vitro* that rs8066588 changes the binding motif of the transcription factor TCF3 (E2A family) (Figure 3B, Table S4), by showing TCF3 preferential binding to the reference C-allele (Figure 3C, Figure S3). Moreover, this variant overlaps an active enhancer element in breast tissue and is associated with robust transcription and a *DNase*I hypersensitive site in mammary cells (Figure 3D, Table S3).

Finally, the candidates rs9891865 and rs9896044, located in *STXBP4* introns, are in strong LD with the daeSNP rs17817901 (r^2^ = 0.8) and the risk lead-variant rs2787486 (r^2^ = 0.75). Both variants are eQTLs to *STXBP4*, are predicted to alter binding motifs of transcription factors expressed in breast and rs9891865 overlaps a *DNase*I hypersensitive site in mammary cells (Figure 3D, Tables S3 and S4) ^41^. Whilst we did not obtain *in-vitro* evidence for protein binding to rs9896044 (data not shown), we could detect differences in the binding of proteins between the two alleles of rs9891865, could not confirm the actual protein involved (Figure S4).

Overall, we found that - variants regulate all three genes in the 17q22 risk locus and that the population variability in AE these genes present is associated with the risk of breast cancer. Furthermore, we identified and validated two *cis*-regulatory variants linked to the AE observed in the three genes. Although the LD is high between the candidate rSNPs, the daeSNPs, and the risk-variant, these variants form four haplotypes (Figure 3E). More importantly, two haplotypes in this region include the C-rs2787486 protective allele: the main haplotype (24.8%) in phase with preferentially expressed alleles in healthy controls, and another less common (5.7%) in phase with the preferentially expressed alleles in cases.

## Discussion

Our work investigated the value of integrating allelic expression (AE) analysis in the validation and characterization of breast cancer risk loci. Importantly, we present a novel approach to assess the association of AE of target genes with risk, suitable to detect association even when multiple *cis*-regulatory variants are involved in a complex risk genetic structure. In the 17q22 risk locus, we identified *TOM1L1, COX11*, and *STXPB4* genes as *cis*-regulated in breast tissue by genetic variants in LD with the GWAS lead SNP in the region. Next, we show that AE of these genes is associated with breast cancer risk with estimated effect sizes ranging from large (detected for rs2628315 in *STXPB4*) to medium (detected for rs17817901 in *COX11/TOM1L1*). Finally, we identified and characterized candidate regulatory variants affecting all target genes, unveiling some of the mechanisms underlying the risk in this locus.

Previous studies have only provided suggestive evidence for genes in this locus to be candidate targets of risk-associated variants. These studies included fine-mapping exercises ^20^, chromatin conformation analysis ^4^, and studies integrating expression data ^5^. Fachal et al provided the most substantial evidence but only scored *STXBP4* and *COX11* to an intermediate level of confidence in the INQUIST algorithm ^5^. Here, we provide compelling evidence that all three genes are candidate targets and that their AE is associated with risk.

We found the most significant association with breast cancer for the AE ratios measured at rs2628315 in an intron of *STXBP4*. This variant is in complete LD with rs2787486, the strongest risk association reported in this locus (OR = 0.92; 95%CI = [0.90; 0.94]; *P* = 8.96 ×10^−15^) ^20^. We found that the G-rs2628315 allele, proxy to the risk C-rs2787486 allele, is 1.5-fold more expressed in cases. Moreover, as *STXBP4* is lowly expressed in breast tissue, this result suggests that the expression of the G-rs2628315 allele is extremely low in healthy tissue, hence increases cancer risk only upon upregulation. Consistently, it suggests an oncogenic role for *STXBP4* in breast cancer. This gene encodes the protein STXBP4 (Syntaxin Binding Protein 4) involved in glucose metabolism, vesicle, and insulin transport. In lung cancer, it binds ΔNp63 (N-terminally truncated isoform of p63), preventing its proteolysis, promoting growth, and blocking cell differentiation, in line with the oncogenic role that our data supports^42,43^.

Moreover, we found an association between AE ratios measured at rs17817901, in a genomic region shared by *TOM1L1* and *COX11*, and breast cancer. Patient samples more often preferentially expressed the reference A-rs17817901 allele, frequently linked to the risk-associated C-rs2787486 allele. Because we found no association for the AE ratios measured at rs9899602, a variant mapping exclusively to the *TOM1L1* sequence, we believe that the association detected at rs17817901 is mostly due to *cis*-regulatory variation acting on *COX11*. rs17817901 is in strong LD with the risk lead-variant rs2787486 (r^2^ = 0.74) but is in even stronger LD (r^2^ = 0.85) with a previously associated variant rs6504950 (OR = 0.95; 95%CI = [0.92; 0.97]; *P* =1.4 × 10^−8^) ^18^. Like the findings for rs2628315 in *STXBP4*, we observed a shift from the controls preferentially expressing the protective G-rs17817901 allele to the patients preferentially expressing the risk A-rs17817901 allele. On average, we found that the patients expressed the risk-associated A-rs17817901 allele 2-fold more than the controls. Additionally, GTEx data shows that *COX11* is a highly expressed gene in breast tissue (higher than *STXBP4*). These data suggest that although *COX11* is already highly expressed, there could be an oncogenic advantage to have its expression further upregulated. *COX11* encodes for a mitochondrial membrane protein crucial for the assembly of an active cytochrome c oxidase complex, which in turn links to the metabolic changes that accompany tumor development ^44,45^, supporting its oncogenic role.

Different tissue types partially share gene expression control, particularly cis-QTLs ^41,46,47^. Our previous work established an overlap between *cis*-regulation of breast cancer genes in breast tissue and blood ^24^. Here verifies whether the risk association found for AE ratios measured in breast tissue were valid in blood, as the testing of this tissue in a future clinical setting greatly facilitates the translation of these results. We found similar effect sizes and directions in the association profiles of AE ratios measured at rs2628315 and rs17817901. This result opens the possibility of carrying out a future genome-wide study for identifying other breast cancer risk-associated daeSNP in blood.

The use of normalized AE ratio distributions also confers robustness to rSNP mapping purposes, as it isolates the effect of *cis*-regulatory variation ^11^. In the case of rs17817901 (*COX11/TOM1L1*), we observed a pronounced shift of the normalized AE ratio distribution from the equimolar AE. This pattern indicates complete LD between the daeSNP and the variant(s) controlling expression and identified two strong candidate rSNPs. The first is rs1781901 itself, predicted to alter the binding of the oncogenic miRNA hsa-miR-194-5p at the 3’ UTR of *COX11*. We established that hsa-miR-194-5p binds preferentially to the alternative C-allele, corresponding to the protective haplotype. Hence, upregulation of the hsa-miR-194-5p expression inhibits the expression of the protective allele, leading to an increased risk of breast cancer. The second candidate rSNP is rs8066588, which overlays an active enhancer associated with robust transcription and a *DNase*I hypersensitive site in mammary cells. We found that the reference C-allele of rs8066588 preferentially bound the transcription factor TCF3, upregulating the expression of the risk-associated allele. Inclusion of these candidate causal rSNPs and risk-daeSNPs in haplotype association studies will further validate our findings.

The approach we present here undoubtedly confirms that AE regulation and *cis*-regulatory variants are involved in the risk of breast cancer in locus 17q22. Using AE ratios as a quantitative continuous variable to compare cases and controls has increased statistical power compared to using discrete variables, as in GWAS. Moreover, although the effect sizes we report using a standardized mean difference method are independent of the sample size, that estimate’s precision depends on it. Therefore, replicate studies using larger sample sizes, especially when reporting novel risk loci, needs to be considered.

AE ratios have been used before to detect association with disease, but not as a quantitative variable to compare cases and control ^16,17^. For example, Valle and colleagues ^16^ calculated normalized ratios and set a cut-off to define samples with and without differential AE, upon which they tested for differences in proportions in the two populations ^16^. However, the different distribution of AE ratios between genes ^24,41^ impedes the establishment of a universal cut-off and statistically lessens the power to compare cases and controls.

Additionally, most of the previously characterized risk loci have shown that rarely a single *cis*-regulatory variant or target gene is involved in risk to disease. When multiple *cis*-regulatory variants are involved, they are also not necessarily in complete/strong LD with each other due to complex genetic architectures. Hence, an additional advantage of our approach is that using AE ratios measured at the candidate target genes, we also detect the cumulative effect of all variants acting on these genes. However, one limitation of our approach is transversal to all studies of AE: only heterozygous individuals for the transcribed variants are informative.

In summary, our work shows that all genes in the risk locus 17q22 are under the control of *cis*-regulatory variants, supports that *STXBP4* and *COX11* are the most likely target genes of the risk-variants identified in previous GWAS, and establishes that AE ratios at two daeSNPs are strongly associated with risk of breast cancer. Our work also unveils the mechanisms underlying disease risk at this locus, indicating an association with a change of preferential expression from the protective GWAS allele (healthy controls) to the risk one (patient samples) in both genes. Overall, we present a novel approach to studying cancer risk, applicable to other complex diseases, using AE ratios as a quantifiable phenotype in case-control studies. This approach facilitates the identification of the risk mechanisms and the target genes that have been challenging in post-GWAS studies and now requires testing in a genome-wide setting to confirm its full potential.

## Data Availability

All data produced will available online at https://github.com/maialab upon publication.

## Description of Supplemental Data

As Supplemental Data include four tables, and four figures.

## Acknowledgments

The authors are deeply appreciative of the contributions made by all sample donors who allow data sets to be created and used for the good of medical care and science. The authors would also like to thank other members of the Functional Genomics of Cancer group at CBMR for helpful discussions and Vitor Morais at UAIC for administrative support.

## Data and Code Availability

Data generated in this study is provided as Supplemental Data and at https://github.com/maialab/17q22AErisk. SNPs associated with breast cancer risk were obtained from the GWAS Catalog website at www.ebi.ac.uk/gwas. SNP data were obtained from the Ensembl database (version 92 and 75) available at www.ensembl.org. Gene and transcript expression, and eQTL data for breast tissue from the GTEx Project (v7) were retrieved from the GTEx Portal at www.gtexportal.org. Other detailed results are available in Supplementary Data and Supplementary Datasets 1 and 2.

