## Supplementary Figures for "Germline Allelic Expression of Genes at 17q22 Locus Associates with Risk of Breast Cancer"

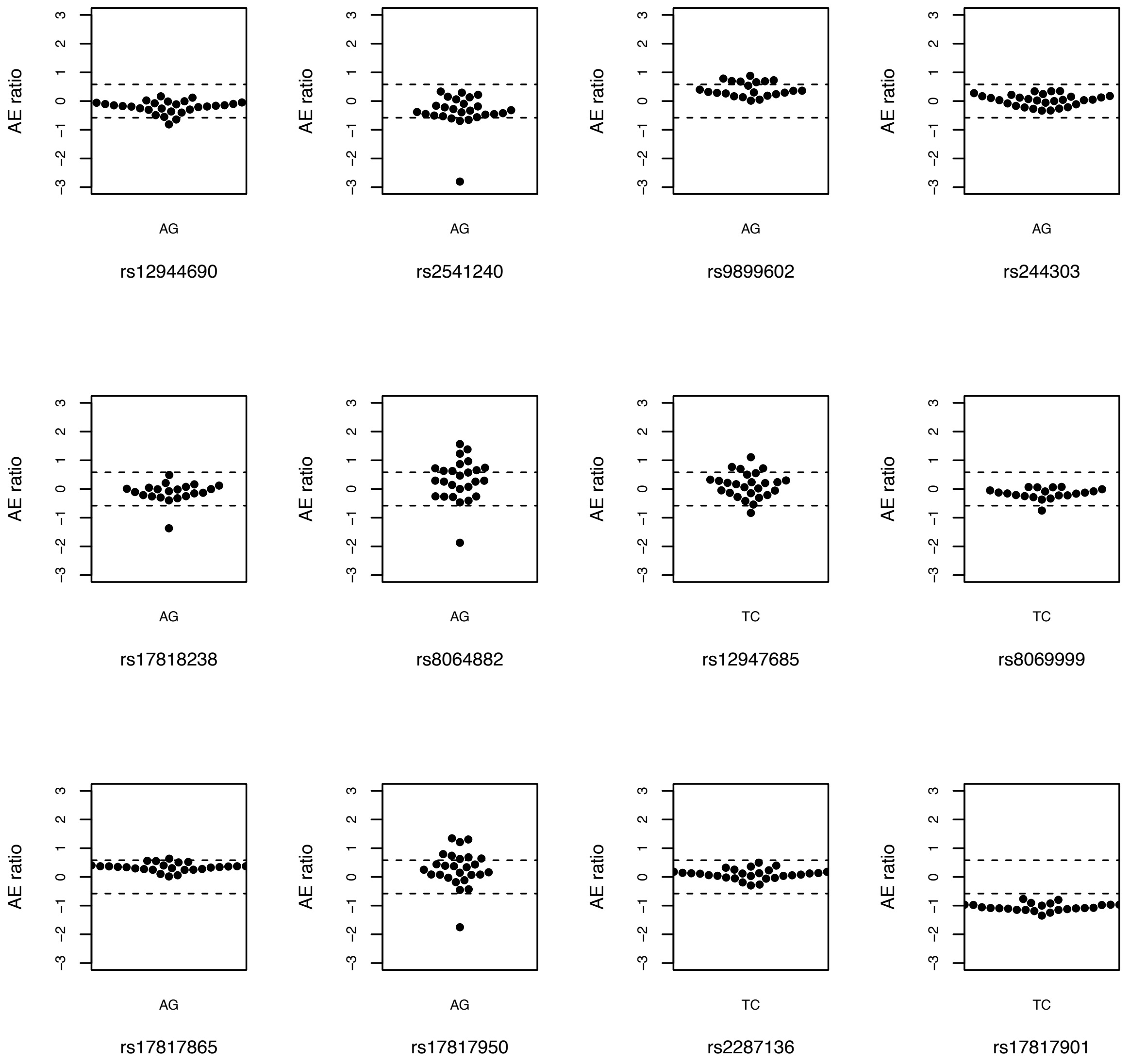

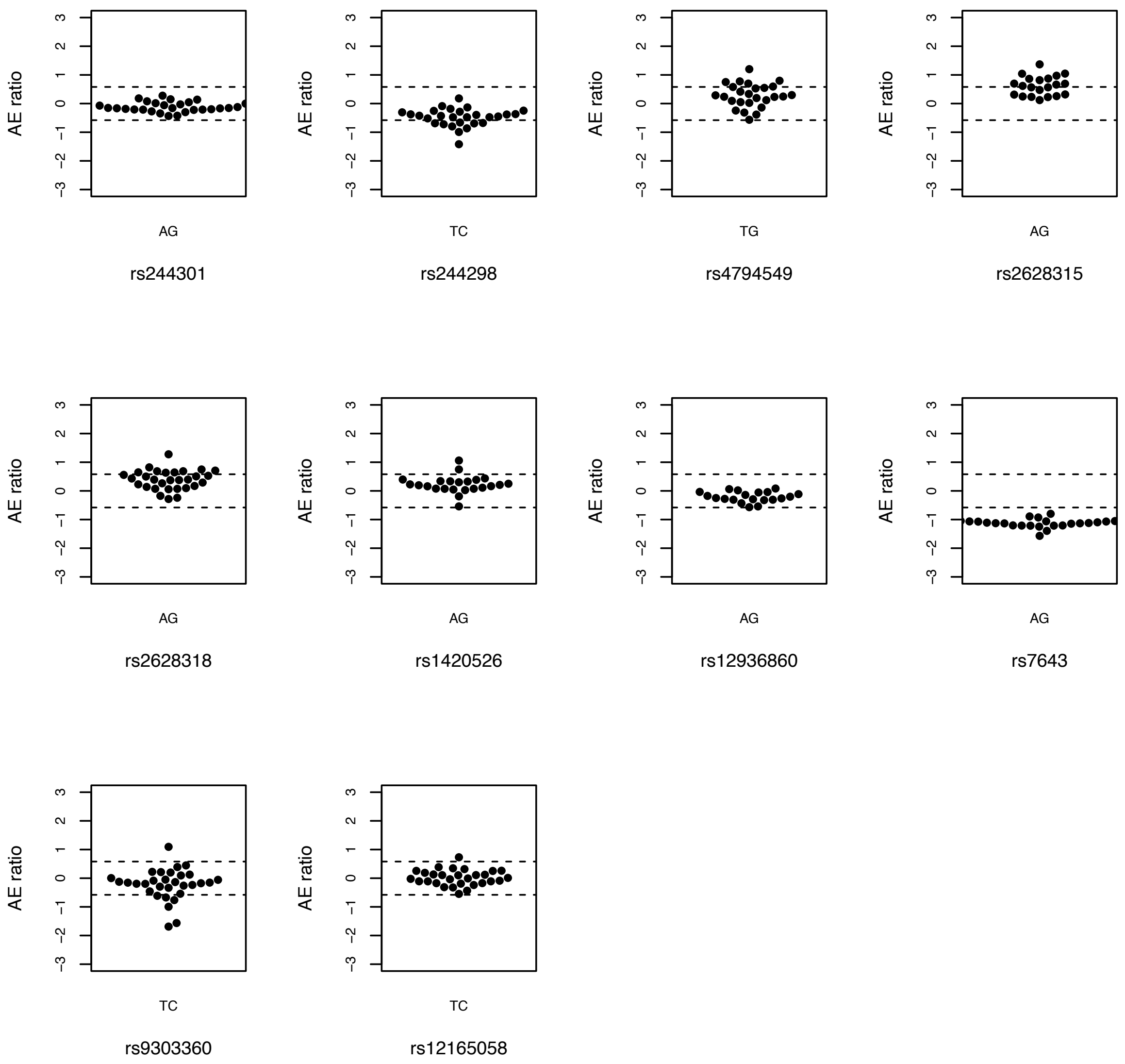


**Figure S1. Allelic expression analysis of 22 variants in the 17q22 risk locus.** Plots of AE ratios (y-axis) calculated for heterozygous individuals (dots) for each variant indicated in the x-axis. The alleles for each variant are indicated below the graphs in the order they were used to calculate the AE ratios: e.g., AB, AE ratio ≈ log2 (allele A/ allele B). Dotted lines indicate 1.5-fold difference between alleles (absolute AE ratio = 0.58).

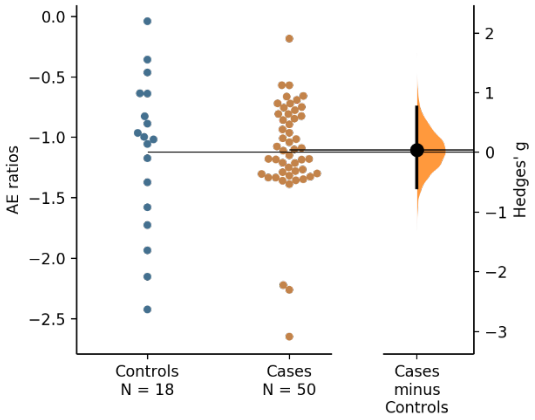


**Figure S2. Case-control study using allelic expression ratios measured at rs9899602 in the 17q22 risk locus in breast tissue samples.** Gardner-Altman estimation plot of Hedges' g between breast cancer cases and controls for allelic expression ratios calculated at rs9899602 (ratio calculated as allele T by allele C) in normal breast tissue. The heterozygous individuals for both groups are plotted on the left axes, with controls displayed in blue and cases in orange. The mean difference is plotted on the floating axes on the right as a bootstrap sampling distribution (bootstrap n = 5000). The mean difference is depicted as a dot, and the 95% confidence interval is indicated by the ends of the vertical error bar.

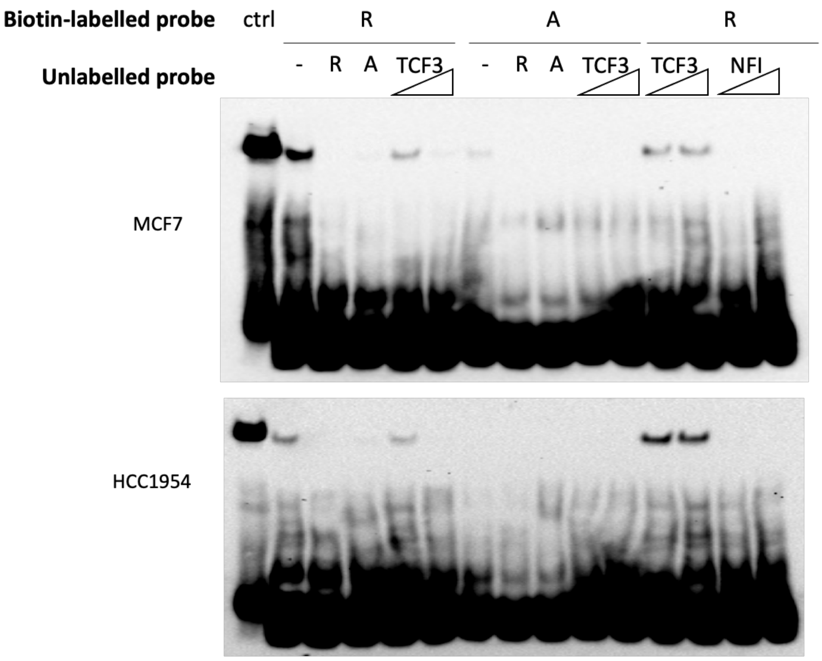


**Figure S3. Complete gel images for EMSA experiments for rs8066588 using protein extracts from MCF-7 and HCC1954 cell lines show preferential binding of the reference C allele** (R – reference, A – alternative alleles). Competition with an oligo of known binding site for TCF3 competes with observed binding, which does not occur with negative control oligo (NFI binding motif).

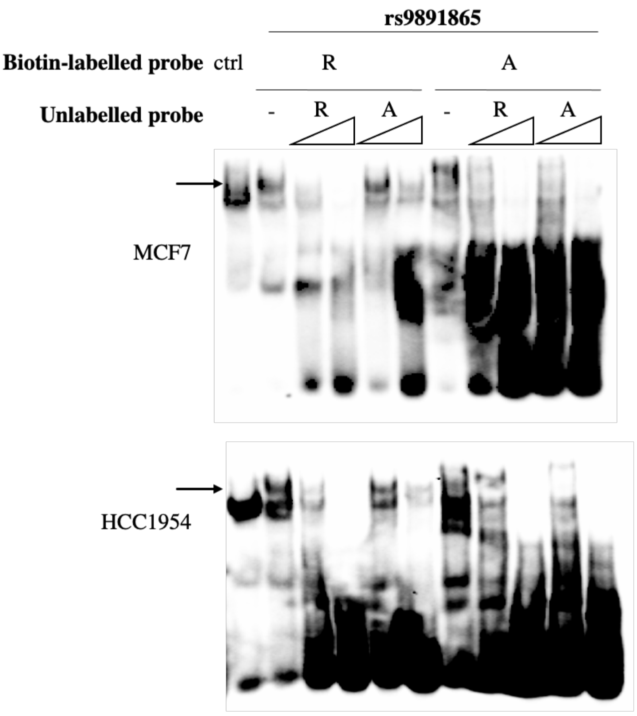


**Figure S4. EMSA experiments for rs9891865 using protein extracts from MCF-7 and HCC1954 cell lines show differential allelic binding** (R – reference, A – alternative alleles). Competition with unlabeled oligos of both alleles show that the high specificity of the binding of the reference allele. Side arrows indicate the position of the reference allele strongest specific band.
