## Supplementary Tables for "Germline Allelic Expression of Genes at 17q22 Locus Associates with Risk of Breast Cancer"

### Table S1. Oligonucleotides used as labelled probes or unlabelled competitors in EMSA analysis, as well as inserts in the reporter assays. The two alleles [reference/alternative] of each SNP are indicated.

| Gene | Method | SNP | Sense strand (5' – 3')<br>Antisense strand (5' – 3') |
|---|---|---|---|
| COX11 | EMSA | rs8066588 | CATTTGCTCAAAACC[C/T]ACCTGTGATTTTCTTCC<br>GGAAGAAAATCACAGGT[G/A]GGTTTTGAGCAAATG |
| | Reporter Assay | rs17817901 | TCGAAGGATCCAATCAGTGTTATCCTGGAACTGTTA[C/T]GCGTCTGTTA[C/T]GCGTCTGTTA[C/T]TCTATTTAATCTTCATTATAGCAGA<br>CTAGTCTGCTATAATGAAGATTAAATAGA[G/A]TAACAGACGC[G/A]TAACAGACGC[G/A]TAACAGTTCCAGGATAACACTGATTGGATCCT |
| STXBP4 | EMSA | rs9896044 | GCATACTTACTTGTTTATTT[C/G]CTATTACAAGAACACTTTCT<br>AGAAAGTGTTCTTGTAATAG[G/C]AAATAAACAAGTAAGTATGC |
| | | rs9891865 | ATTACTGGAATCCCTGTTGG[C/T]TGAGGGCCTTGCAAGTTTGT<br>ACAAACTTGCAAGGCCCTCA[G/A]CCAACAGGGATTCCAGTAAT |

### Table S2. Summary statistics for differential allelic expression analysis in the 17q22 risk locus.

| SNP | Position (hg38) | Gene | Ref Allele | Alt Allele | MAF | LD with rs2787486 (r2) | P-value | p.adjust | mean | CIinf | CIsup |
|---|---|---|---|---|---|---|---|---|---|---|---|
| rs9899602 | 54909547 | TOM1L1 | T | C | 0.29 | 0.527061 | 1.88e-07 | 6.83e-07 | 0.410 | 0.298 | 0.522 |
| rs9303360 | 54924095 | TOM1L1 | T | C | 0.42 | 0.192934 | 0.027 | 0.036 | -0.334 | -0.629 | -0.040 |
| rs12165058 | 54944688 | TOM1L1 | T | C | 0.46 | 0.166612 | 0.768 | 0.796 | 0.015 | -0.089 | 0.119 |
| rs12944690 | 54952253 | TOM1L1/COX11 | A | G | 0.46 | 0.163259 | 4.21e-05 | 1.22e-04 | -0.199 | -0.284 | -0.115 |
| rs12936860 | 54952865 | TOM1L1/COX11 | G | A | 0.27 | 0.570168 | 6.97e-05 | 1.84e-04 | -0.209 | -0.295 | -0.122 |
| rs17817865 | 54955064 | TOM1L1/COX11 | G | A | 0.27 | 0.094121 | 1.22e-13 | 1.18e-12 | 0.350 | 0.296 | 0.404 |
| rs2287136 | 54960721 | TOM1L1/COX11 | G | A | 0.27 | 0.093248 | 6.35e-03 | 9.21e-03 | 0.098 | 0.030 | 0.166 |
| rs17817901 | 54961384 | TOM1L1/COX11 | A | G | 0.27 | 0.570168 | 1.66e-22 | 2.41e-21 | -1.050 | -1.107 | -0.995 |
| rs7643 | 54962122 | TOM1L1/COX11 | A | G | 0.46 | 0.16787 | 1.71e-26 | 4.95e-25 | -1.110 | -1.166 | -1.054 |

| | | | | | | | | | | | |
|---|---|---|---|---|---|---|---|---|---|---|---|
| rs2541240 | 54965511 | COX11 | G | A | 0.33 | 0.14134 | 4.21e-03 | 6.42e-03 | -0.359 | -0.594 | -0.124 |
| rs17817950 | 54967964 | COX11/STXBP4 | G | A | 0.27 | 0.116478 | 0.033 | 0.042 | 0.277 | 0.024 | 0.531 |
| rs12947685 | 54968158 | COX11/STXBP4 | T | C | 0.27 | 0.116478 | 0.141 | 0.158 | 0.131 | -0.047 | 0.309 |
| rs8064882 | 54968848 | COX11/STXBP4 | C | T | 0.27 | 0.11699 | 0.037 | 0.045 | 0.304 | 0.019 | 0.589 |
| rs4794549 | 54969365 | COX11/STXBP4 | C | A | 0.29 | 0.130241 | 1.79e-03 | 3.16e-03 | 0.282 | 0.116 | 0.449 |
| rs1420526 | 55052047 | STXBP4 | T | C | 0.26 | 0 | 1.69e-03 | 3.16e-03 | 0.227 | 0.095 | 0.358 |
| rs17818238 | 55068243 | STXBP4 | G | A | 0.21 | 0.131223 | 0.136 | 0.157 | -0.120 | -0.280 | 0.041 |
| rs8069999 | 55081131 | STXBP4 | G | A | 0.19 | 0.130523 | 1.57e-03 | 3.16e-03 | -0.159 | -0.249 | -0.069 |
| rs2628318 | 55132951 | STXBP4 | G | A | 0.42 | 0.288655 | 8.91e-07 | 2.87e-06 | 0.387 | 0.259 | 0.514 |
| rs2628315 | 55149261 | STXBP4 | G | A | 0.28 | 0.985466 | 5.18e-08 | 2.15e-07 | 0.617 | 0.464 | 0.770 |
| rs244303 | 55161367 | STXBP4 | A | G | 0.41 | 0.267585 | 0.467 | 0.502 | 0.028 | -0.050 | 0.105 |
| rs244301 | 55161779 | STXBP4 | T | C | 0.41 | 0.265277 | 1.96e-03 | 3.16e-03 | -0.115 | -0.183 | -0.046 |
| rs244298 | 55164026 | STXBP4 | T | C | 0.41 | 0.267585 | 7.41e-09 | 3.58e-08 | -0.486 | -0.607 | -0.365 |
| rs171512 | 55196232 | downstream STXBP4 | G | T | 0.35 | 0 | 1.02e-03 | 2.27e-03 | 0.306 | 0.135 | 0.478 |
| rs2287223 | 55315491 | HLF | G | A | 0.37 | 0 | 3.27e-04 | 7.91e-04 | 0.172 | 0.088 | 0.256 |
| rs17746075 | 55321098 | HLF | T | G | 0.15 | 0 | 1.92e-03 | 3.16e-03 | 0.183 | 0.081 | 0.285 |
| rs11547827 | 55393101 | MMD | C | T | 0.27 | 0 | 2.25e-12 | 1.63e-11 | -0.417 | -0.481 | -0.353 |
| rs4476230 | 55393744 | MMD | T | G | 0.39 | 0 | 1.17e-09 | 6.77e-09 | -0.273 | -0.335 | -0.210 |
| rs4450463 | 55404443 | MMD | C | T | 0.034 | 0 | 0.012 | 0.0174 | 0.333 | 0.120 | 0.546 |
| rs4794600 | 55546472 | upstream SMIM36 | G | A | 0.32 | 0 | 0.859 | 0.859 | 0.024 | -0.256 | 0.305 |

**Legend:**

MAF – minor allele frequency in the 1000GENOMES:phase_3:EUR population; P-value – p-value of a two-sided t-test for mean equal to zero; p.adjust – p-value adjusted for multiple testing with fdr correction; CIinf and CIsup – Inferior and superior limits of the 95% confidence interval (CI) of the mean.

**Table S3. Functional annotations of 106 candidate regulatory variants in the 17q22 risk locus, retrieved from Haploreg v4.1 and RegulomeDB.**

Please see Esteves_TableS3.csv at:

https://ualg365-my.sharepoint.com/:x:/g/personal/atmaia_ualg_pt/EX-IHyw8EM9PkYhC6NsQ9o4BBmISk8B-W7AUFy98zIuUdA?e=er0Dd8

**Legend:**

Chr – chromosome; pos_hg38 – genomic position according to hg38; rsID - variant ID in dbSNP build 141; ref – reference allele; alt - alternative allele; AFR, AMR, ASN, EUR –

minor allele frequency in continental populations (AFR, AMR, ASN, EUR); Chromatin_States Imputed - chromatin state segmentations (15-state and 25-state) from the Roadmap Epigenomics Project; Chromatin_Marks - chromatin mark ChIP-seq tracks (gappedPeak calls) from the Roadmap Epigenomics Project; DNAse - DNase tracks (narrowPeak calls) from the Roadmap Epigenomics Project; ChIP-Seq - SPP narrow peaks called by the ENCODE project; eQTL – eQTLs identified mainly in the GTEx analysis V6 and the GEUVADIS analysis; GWAS – GWAS NHGRI Catalog data; GRASP – data from GRASP Build 2.0.0.0; PWM Motifs – significant position weight matrices (PWMs) for known protein motifs; GENCODE_id and GENCODE_name – Gene ID and name in GENCODE version 13; RefSeq_id and RefSeq_name – Gene ID and name in the RefSeq NCBI Reference Sequence Database.

**Table S4. Regulatory motifs altered by candidate rSNPs, data from Haploreg v4.1.** Position weight matrices (PWMs) were calculated as described in the documentation of Haploreg v4.1 (.../haploreg/documentation_v4.1.html). The change in log-odds (LOD) score for reference and alternative alleles is presented for motifs which passed significant PWM calculation for both alleles and overlapped the variant nucleotide.

| Candidate SNP | Position Weight Matrix ID | Strand | Ref LOD Score | Alt LOD Score | Allelic Difference |
|---|---|---|---|---|---|
| **rs9891865** | E2A_5 | - | 11.7 | 3.8 | 7.9 |
| | Spz1_1 | - | 11.9 | 0.1 | 11.8 |
| | ZEB1_disc1 | - | 12.7 | 10.7 | 2 |
| **rs8066588** | NF-I_2 | + | 11.9 | 5.8 | 6.1 |
| | SMC3_disc2 | - | -6.7 | 5.2 | -11.9 |
| | TCF12_disc2 | - | 12.1 | 0.1 | 12 |
| | THAP1_disc1 | + | 3.4 | 4.6 | -1.2 |
| **rs9896044** | ELF1_known1 | - | 15.3 | 3.4 | 11.9 |
| | Fox | + | 2.1 | 13 | -10.9 |
| | Foxa_known1 | + | 0.9 | 12.2 | -11.3 |
| | Mef2_disc2 | - | 14.9 | 2.9 | 12 |
| | Pou3f2_1 | - | 12.4 | 1.7 | 10.7 |
| | TEF | + | -0.5 | 11.1 | -11.6 |
